## Supplemental Data for "Performance of SARS-CoV-2 Serology tests: Are they good enough?"

**Supplemental Table:** Characteristics of the immunoassays evaluated, as provided by the manufacturers.

| **Assay and manufacturer** | **Analyser used**  **(if any)** | **Viral target and antibody type** | **Manufacturer's thresholds (R)** |
| --- | --- | --- | --- |
| **Epitope Diagnostics Ltd.**  **SARS-CoV-2 ELISA** | Agility (Dynex) | Nucleocapsid protein, IgG | R=OD_sample_/1.1x(NC)+0.18) |
|  |  |  | NC=mean of negative control OD |
|  |  |  | R<1 : negative |
|  |  |  | R≥1 : positive |
| **EuroImmun**  **SARS-CoV-2 ELISA** | Manual | Spike protein S1 IgG | OD_sample_/OD_cal_ |
|  |  |  | R<0.8 : negative |
|  |  |  | 0.8≤R<1.1: equivocal |
|  |  |  | R≥1 : positive |
| **DiaSorin Laison**  **SARS-CoV-2 S1/S2 IgG Immunoassay** | Liaison XL | Spike protein S1/S2 IgG | 3-points calibration curve |
|  |  |  | R<12 AU/mL : negative |
|  |  |  | 12≤ R < 15 AU/mL : equivocal |
|  |  |  | ≥15 : positive |
| **Abbott Diagnostics**  **SARS-CoV-2 Immunoassay** | Alinity | Nucleocapsid protein, IgG | OD_sample_/OD_cal_ |
|  |  |  | R<1.4 : negative |
|  |  |  | R≥1.4 : positive |
| **Healgen**  **COVID-19 IgG/IgM Rapid Test** | Cassette provided | IgG/IgM - specificity of antigen not given | IgG band present = G positive |
|  |  |  | IgM band present = M positive |
